# Associations between use of aspirin and MRI-derived liver fat and fibroinflammation

**DOI:** 10.64898/2025.12.23.25342902

**Authors:** Qi Feng, Pinelopi Manousou, Chioma N Izzi-Engbeaya, Jun Liu, Mark Woodward

**Author notes:** Correspondence to: Dr. Qi Feng, the George Institute for Global Health (UK), Scale Space, Imperial College London, 58 Wood Lane, London W12 7RZ UK. No funding was received for this work. The authors declared no competing interests for this work.

## Abstract

**Background:** The effects of aspirin on hepatic steatosis and fibroinflammation are unclear. The study aimed to examine the association between aspirin use and liver MRI-derived liver fat and corrected T1 (cT1).

**Methods:** We used UK Biobank imaging cohort data. Aspirin use was self-reported at baseline and imaging assessment, and the main exposures were aspirin use at imaging assessment and longitudinal aspirin use patterns (never users, initiators, discontinuers, vs. persistent users). Outcomes were MRI-derived liver fat (%) and cT1 (ms). Multivariable adjustment analyses and inverse probability of treatment weighting (IPTW) analyses were performed, accounting for demographic, lifestyle and clinical factors.

**Results:** We included 36413 participants (age 64.6 years, 51.4% female). Aspirin use at imaging assessment was associated with lower liver fat (-0.35 (95% CI: -0.51, - 0.20)) and slightly higher cT1 (5.13 (95% CI: 3.23, 7.03)). Analyses on longitudinal aspirin use pattern showed that compared to never users, initiators and persistent users showed lower liver fat (-0.48 (-0.69, -0.28) and (-0.24 (-0.45, -0.02)) and higher cT1 (2.94 (0.38, 5.49) and 8.31 (5.65, 10.97)). IPTW analyses showed consistent results.

**Conclusion:** In this large population-based cohort, aspirin use was linked to reduced liver fat, but a small, clinically insignificant (i.e. <80ms) increase in cT1. These findings suggest aspirin may mitigate steatosis through metabolic pathways but does not necessarily rapidly reverse fibroinflammatory injury.

## Introduction

Aspirin is among the most widely used medications worldwide, primarily for primary and secondary prevention in people with CVD [1–3]. Steatotic liver disease (SLD) affects over one third of the global population, making it the most common chronic liver disease [4]. It is defined by the excessive accumulation of fat at the liver [5,6]. Beyond its hepatic manifestation, liver steatosis is strongly linked to cardiometabolic risk factors, and is associated with various extrahepatic diseases, including cardiovascular disease (CVD), cancers, and chronic kidney disease [7–9].

The potential effects of aspirin on liver steatosis have been the focus of increasing research, but the findings have been inconsistent. Early cross-sectional studies suggested associations between aspirin use and lower prevalence of SLD [10], as well as less severe histologic features and lower risk of progression to advanced fibrosis in SLD populations [11,12]. A recent review further concluded that most studies supported a protective effect of aspirin on SLD prevalence and progression [13]. In contrast, Huang et al [14] reported a positive association between aspirin use and MASLD risk, while a US study found no association between aspirin and liver fibrosis measures in people with myocardial infarction [15]. Evidence from randomized controlled trials is limited, with one small trial reporting that low-dose aspirin reduced liver fat content in 80 non-cirrhotic MASLD patients [16].

These discrepancies may reflect key limitations of prior research. Firstly, most existing studies relied on clinical diagnoses of SLD from electronic health records, which is prone to underestimation of SLD prevalence and incidence [17]. Few studies have examined the associations with accurate imaging-based measures of liver fat percentage and liver disease activity in general populations. Secondly, many previous studies assessed aspirin exposure only cross-sectionally, without accounting for longitudinal pattern of aspirin use, whilst, thirdly, their sample sizes have often been modest.

To address these gaps, the present study aimed to investigate the associations of aspirin use and longitudinal use pattern, with imaging-derived liver fat and liver disease activity in a large set of UK Biobank (UKB) participants.

## Material and Methods

The UKB cohort study comprises approximately half million individuals recruited between 2006 and 2010. At baseline assessment, participants completed questionnaire on sociodemographic background, lifestyle, and medical history. Biological samples of blood, urine and saliva were also collected [18]. Since 2014, participants have been invited to attend an imaging visit, which included magnetic resonance imaging (MRI) scans of the brain, heart and abdominal organs. As of September 2025, liver MRI data are available for around 40000 participants [19]. Baseline data were updated during imaging visits; however, biochemistry data were not available at imaging visits.

We excluded participants who did not have liver MRI data, had withdrawn consent, had chronic liver conditions other than SLD at baseline or at imaging assessment (e.g., viral hepatitis, liver fibrosis, liver cirrhosis, hepatocellular carcinoma, hemochromatosis, Wilson’s disease, biliary cirrhosis, autoimmune hepatitis, primary sclerosing cholangitis, toxic liver disease and Budd-Chiari syndrome), or had missing data in key variables (sociodemographic, lifestyle, cardiometabolic risk factors).

### Exposure and outcome

At both UKB baseline and imaging visits, participants reported whether they regularly used aspirin (yes/no), defined as taking it on most days during the past four weeks. The primary exposure was aspirin use at imaging visit. We assessed longitudinal pattern of aspirin use based on the responses at baseline and imaging visits, classified as never users (no->no), initiators (no->yes), discontinuers (yes->no) and persistent users (yes->yes).

Liver MRI scans were acquired using a Siemens MAGNETOM Aera1.5T scanner (Syngo MR D13) and the LiverMultiScan protocol from Perspectum Ltd (UK) [20]. Liver fat (%) was quantified using proton density fat fraction (PDFF). Liver steatosis was defined as liver fat ≥ 5% [21]. Liver iron corrected T1 (cT1, measured in milliseconds (ms)) was also derived from liver MRI, which is highly correlated to liver inflammation and fibrosis [22,23]. MRI T1 relaxation time reflects extracellular fluid, which is characteristic of fibrosis and inflammation. The presence of iron, which can be determined from T2* maps, has an opposing effect. Combining T2* and T1 values can correct for this opposing effect, from which cT1 is derived. Higher cT1 values are associated with histological liver inflammation and fibrosis, although their relative contributions to the score are unclear [24]. Liver fat content and cT1 have been recommended outcomes for assessing treatment-induced histological improvements in people with metabolic dysfunction associated steatohepatitis [25].

### Covariates

Socioeconomic status was measured using the Townsend Deprivation Index, a postcode-based social deprivation score. Self-reported educational attainment was categorised as below secondary, lower secondary, upper secondary, vocational training, and higher education. Self-recorded ethnic background was classified into White or other.

Smoking status was self-reported as current, previous or never smokers. Alcohol consumption was assessed via self-reported intake of various alcoholic drinks; the consumption was summed up to derive average daily alcohol consumption (g/d).

Physical activity level was measured with the International Physical Activity Questionnaire, and individuals were categorized into low, moderate and high levels, based on the frequency, duration and intensity of their physical activities.

Systolic and diastolic blood pressures were measured twice by trained staff, and the average was used for analysis. Blood biochemistry markers were measured at a central laboratory [21]. Diabetes was defined as glycated haemoglobin (HbA1c) ≥39 mmol/mol and/or diagnosis of type 2 diabetes and/or on treatment for type 2 diabetes. Hypertension was defined as systolic blood pressure (BP) ≥130 and/or diastolic BP ≥ 85 mmHg and/or on antihypertensive drug treatment or diagnosis of hypertension. Body mass index (BMI) was calculated as weight (kg) divided by height squared (m^2^). High triglycerides (TG) were defined as plasma TG ≥ 1.70 mmol/L and/or on lipid lowering treatment. Low high-density lipoprotein (HDL) cholesterol was defined as HDL-cholesterol ≤ 1.0 mmol/L (male) ( ≤ 1.3 mmol/L (female)) and/or on lipid lowering treatment. Participants also reported whether they have had any existing cardiovascular disease (CVD), including angina, stroke and myocardial infarction. Since liver fat was not measured at baseline, we calculated fatty liver index (FLI) as a surrogate for liver fat at baseline assessment; FLI was calculated based on BMI, waist circumference, TG level and gamma-glutamyl transpeptidase level [26].

### Statistical *analysis*

Participants’ characteristics at imaging visit were summarised stratified by aspirin use status at imaging visit and by longitudinal pattern of aspirin use.

We fitted linear regression to examine the associations between aspirin use and longitudinal aspirin use pattern with liver fat and cT1, with beta and 95% confidence interval (CI) as effect measure. Logistic regression was fitted for liver steatosis, with odds ratio (OR) and 95%CI as effect measure. The models were adjusted for age, sex, ethnicity, Townsend Deprivation Index (in fifths), education, smoking, physical activity, daily alcohol consumption, BMI, diabetes, hypertension, high TG, low HDL, existing CVD and baseline FLI. The associations of aspirin use at baseline visit was adjusted for covariates measured at baseline, and the associations of aspirin use at imaging visit was adjusted for covariates measured at imaging visit.

We conducted subgroup analyses stratified by sex, age group (< 65, ≥ 65), physical activity, alcohol drinking level (<20/30, 20/30-50/60, ≥ 50/60 g/d for female/male), BMI groups (< 25, 25-30, ≥ 30 kg/m^2^), existing CVD, diabetes, hypertension, high TG, low HDL, and baseline FLI (<60, ≥ 60).

To further address confounding bias, we performed inverse probability of treatment weight (IPTW) analysis. First, we estimated propensity score using logistic regression models for aspirin use, adjusted for the covariates mentioned above. Second, we calculated IPTW based on the marginal probability of the observed exposure divided by the predicted propensity score. Third, we examined the overlap of propensity score distributions between groups to ensure adequate overlap. Fourth, we ran a weighted regression model of the outcome on aspirin use, using the calculated IPTW as weights. We fitted multinominal regression to generate propensity score for longitudinal aspirin use patterns. For sensitivity analysis, IPTW were truncated at the 0.1% and 99.9% percentiles to limit the influence of extreme values.

## Results

This study included 36413 participants (mean age 64.6 years, 48.6% males; figure 1). At imaging assessment, 3530 (9.7%) participants reported regular aspirin use. Compared to non-users, aspirin users were more likely to be males, older, socioeconomically deprived, less educated, former or current smokers, and physically inactive, and to consume more alcohol. They also had higher baseline FLI, BMI, and systolic blood pressure, and higher prevalence of hypertension, diabetes, dyslipidaemia, as well as existing CVD. (table 1)

**Figure 1:**
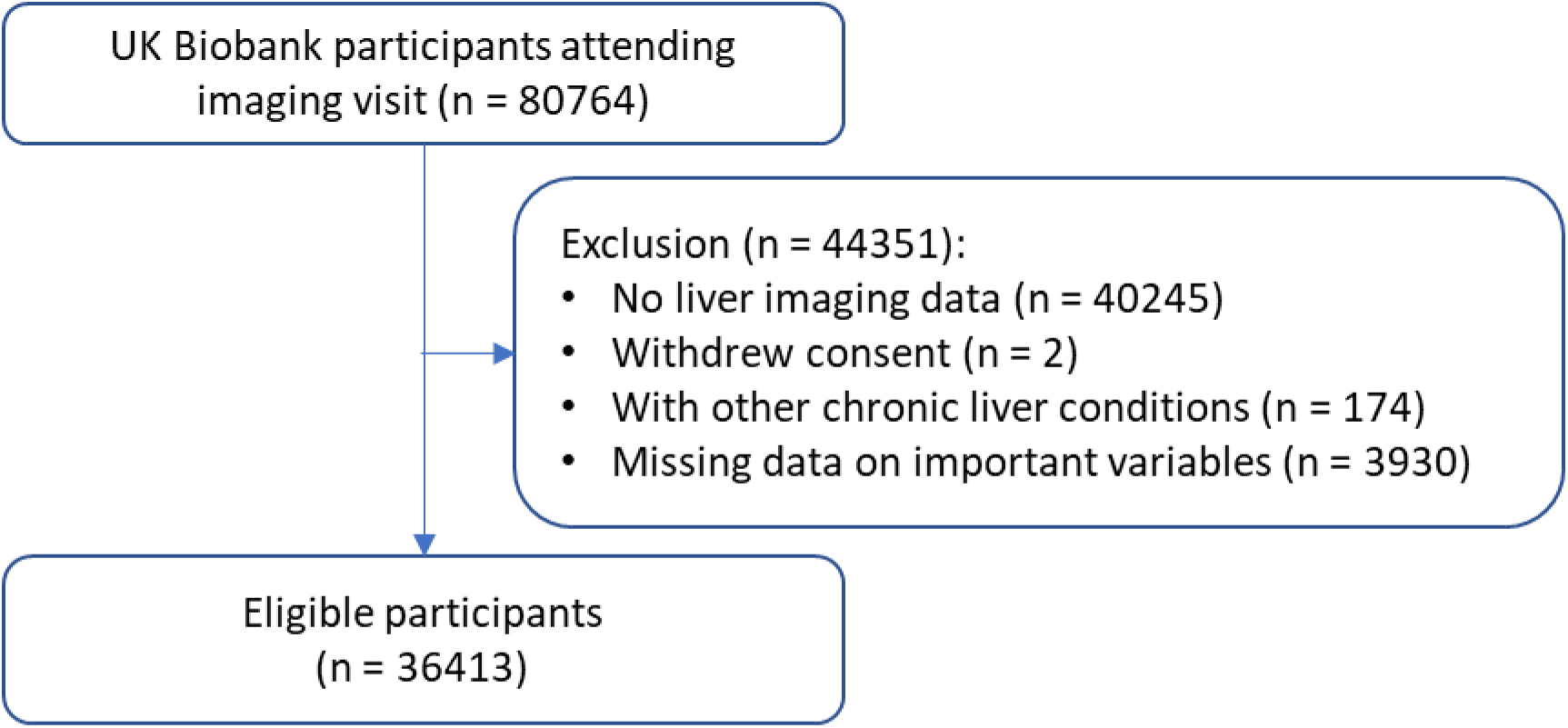
Flowchart of participant selection

**Table 1:** Participants’ characteristics at imaging visit, stratified by aspirin use at imaging visit.

|  | Overall<br>n = 36413<br>(100.0%) | Non-user<br>n = 32883<br>(90.3%) | Aspirin user<br>n = 3530<br>(9.7%) | P value |
| --- | --- | --- | --- | --- |
| Sex, male | 17683 (48.6%) | 15277 (46.5%) | 2406 (68.2%) | < 0.01 |
| Age, years | 64.6 (7.7) | 64.2 (7.6) | 67.4 (7.2) | < 0.01 |
| Townsend Deprivation Index |  |  |  | 0.14 |
| 1st fifth (least deprived) | 7283 (20%) | 6618 (20.1%) | 665 (18.8%) |  |
| 5th fifth (most deprived) | 7239 (19.9%) | 6497 (19.8%) | 742 (21%) |  |
| Education, higher education | 17728 (48.7%) | 16159 (49.1%) | 1569 (44.4%) | < 0.01 |
| Ethnicity, White | 35267 (96.9%) | 31869 (96.9%) | 3398 (96.3%) | 0.04 |
| Smoking, never | 22788 (62.6%) | 20902 (63.6%) | 1886 (53.4%) | < 0.01 |
|  | 10.3 (2.7, |  |  |  |
| Alcohol drinking, g/d* | 20.6) | 10.3 (2.7, 20.1) | 11.4 (2.8, 22.8) | < 0.01 |
| Physical activity, high | 14519 (39.9%) | 13193 (40.1%) | 1326 (37.6%) | 0.02 |
| Body mass index, kg/m<sup>2</sup> | 26.5 (4.3) | 26.4 (4.3) | 27.4 (4.3) | < 0.01 |
| Waist circumference, cm | 88.2 (12.5) | 87.8 (12.5) | 92.5 (12.1) | < 0.01 |
| Systolic blood pressure, mmHg | 138.7 (18.5) | 138.5 (18.5) | 141.0 (18.5) | < 0.01 |
| Diastolic blood pressure, mmHg | 78.7 (10.0) | 78.7 (10.0) | 78.1 (10.1) | < 0.01 |
|  |  |  | 51.1 | < 0.01 |
| Fatty liver index baseline | 41.1 (29.3) | 40.0 (29.1) | (29.1) |  |
| Hypertension | 29439 (80.8%) | 26256 (79.8%) | 3183 (90.2%) | < 0.01 |
| Diabetes | 5045 (13.9%) | 4181 (12.7%) | 864 (24.5%) | < 0.01 |
| High triglycerides | 14319 (39.3%) | 12529 (38.1%) | 1790 (50.7%) | < 0.01 |
| Low HDL-cholesterol | 11707 (32.2%) | 10256 (31.2%) | 1451 (41.1%) | < 0.01 |
| Existing cardiovascular disease | 422 (1.2%) | 147 (0.4%) | 275 (7.8%) | < 0.01 |
| Liver steatosis | 10130 (27.8%) | 8970 (27.3%) | 1160 (32.9%) | < 0.01 |
| Liver fat, % | 4.9 (4.9) | 4.9 (4.9) | 5.4 (5.2) | < 0.01 |
| Liver fat, %* | 3.1 (2.2, 5.5) | 3.0 (2.2, 5.4) | 3.4 (2.4, 6.5) | < 0.01 |
| Liver cT1, ms | 700.6 (54.9) | 699.0 (54.5) | 715.7 (56.0) | < 0.01 |

HDL: high density lipoprotein. Diabetes was defined as glycated haemoglobin (HbA1c) $\geq 39$
mmol/mol and/or diagnosis of type 2 diabetes and/or on treatment for type 2 diabetes.
Hypertension was defined as systolic blood pressure (BP) $\geq 130$ and/or diastolic BP $\geq 85$
mmHg and/or on antihypertensive drug treatment or diagnosis of hypertension. High TG was
defined as plasma TG $\geq 1.70$ mmol/L and/or on lipid lowering treatment. Low HDL cholesterol
was defined as HDL-cholesterol $\leq 1.0$ mmol/L (male) ( $\leq 1.3$ mmol/L (female)) and/or on lipid
lowering treatment. \*: showing median (interquartile interval).

By longitudinal aspirin use patterns, 30466 (83.7%) participants as never users, 1772 (4.9%) initiators, 2417 (6.6%) discontinuers, and 1758 (4.8%) persistent users. Across these groups, there was a graded increase in the likelihood of being males, older age, socioeconomically deprived, less educated, smokers, and having higher alcohol drinking, BMI, dyslipidemia, and existing CVD. Discontinuers had slightly higher prevalence of diabetes than initiators. (Supplementary table 1)

In multivariable-adjusted analyses, aspirin use at baseline visit showed null associations with liver fat or cT1. However, aspirin use at imaging visit was inversely associated with liver fat (beta (95% CI): -0.35 (-0.51, -0.20)), and odds of liver steatosis (OR (95%CI): 0.83 (0.75, 0.90)). IPTW analyses yielded similarly negative associations with liver fat (-0.25 (-0.44, -0.05)) and liver steatosis (0.88 (0.80, 0.96)). By contrast, aspirin use was associated positively with higher liver cT1, with beta of 5.13 (3.23, 7.03) and 5.06 (2.41, 7.71) for multivariable adjustment analysis and IPTW analysis. (table 2)

**Table 2:** Association between aspirin use at baseline and imaging visits and liver fat and cT1(at imaging visit)

|  | Aspirin use at<br>baseline visit | Aspirin use at<br>imaging visit |  |
| --- | --- | --- | --- |
|  | Multivariable<br>adjustment | Multivariable<br>adjustment | IPTW |
| Liver fat, % | -0.05 (-0.22, 0.12) | -0.35 (-0.51, -<br>0.20) | -0.25 (-0.44, -0.05) |
| Liver<br>steatosis* | 0.99 (0.90, 1.08) | 0.83 (0.75, 0.90) | 0.88 (0.80, 0.96) |
| Liver cT1, ms | 6.29 (4.20, 8.39) | 5.13 (3.23, 7.03) | 5.06 (2.41, 7.71) |

Multivariable adjustment model included age, sex, deprivation index, ethnicity, physical activity, smoking, daily alcohol consumption, diabetes, hypertension, body mass index, high triglycerides, low high-density lipoprotein, existing cardiovascular disease, and baseline fatty liver index. For association of aspirin use at baseline visit, the covariates were measured at baseline visit. for association of aspirin use at imaging visit, the covariates were measured at imaging visit. \*: showing odds ratio (95CI) for liver steatosis, and beta (95%CI) for liver fat and cT1.

Comparing with never users in multivariable adjustment analysis, initiators and persistent users demonstrated lower liver fat (-0.48 (-0.68, -0.27) and -0.24 (-0.45, - 0.03), respectively), while discontinuers showed non-significant association (-0.10 (-0.27, 0.08)). Similarly, initiators and persistent users showed lower prevalence of liver steatosis (0.77 (0.68, 0.87), 0.88 (0.78, 0.99)), while discontinuers showed non-significant association (0.96 (0.87, 1.07)). Initiators, discontinuers and persistent users progressively showed higher cT1 (2.99 (0.44, 5.55), 4.12 (1.91, 6.33), 8.35 (5.70, 11.01)). IPTW analyses showed similar results. Compared to never users, initiators had lower liver fat (-0.30 (-0.55, -0.06)), persistent users marginally non-significant associations (-0.34 (-0.68, 0.01)), while discontinuers showed null associations (-0.12 (-0.35, 0.10)). Initiators and persistent users showed lower odds of liver steatosis (0.87 (0.77, 0.98), and 0.83 (0.71, 0.98)), while discontinuers showed non-significant associations (0.93 (0.84, 1.03)). Compared with never users, initiators, discontinuers and persistent users progressively showed higher cT1 (3.01 (-0.23, 6.25), 3.11 (0.08, 6.14), 7.01 (2.02, 12.00)), in IPTW analysis. (Figure 2)

**Figure 2:**
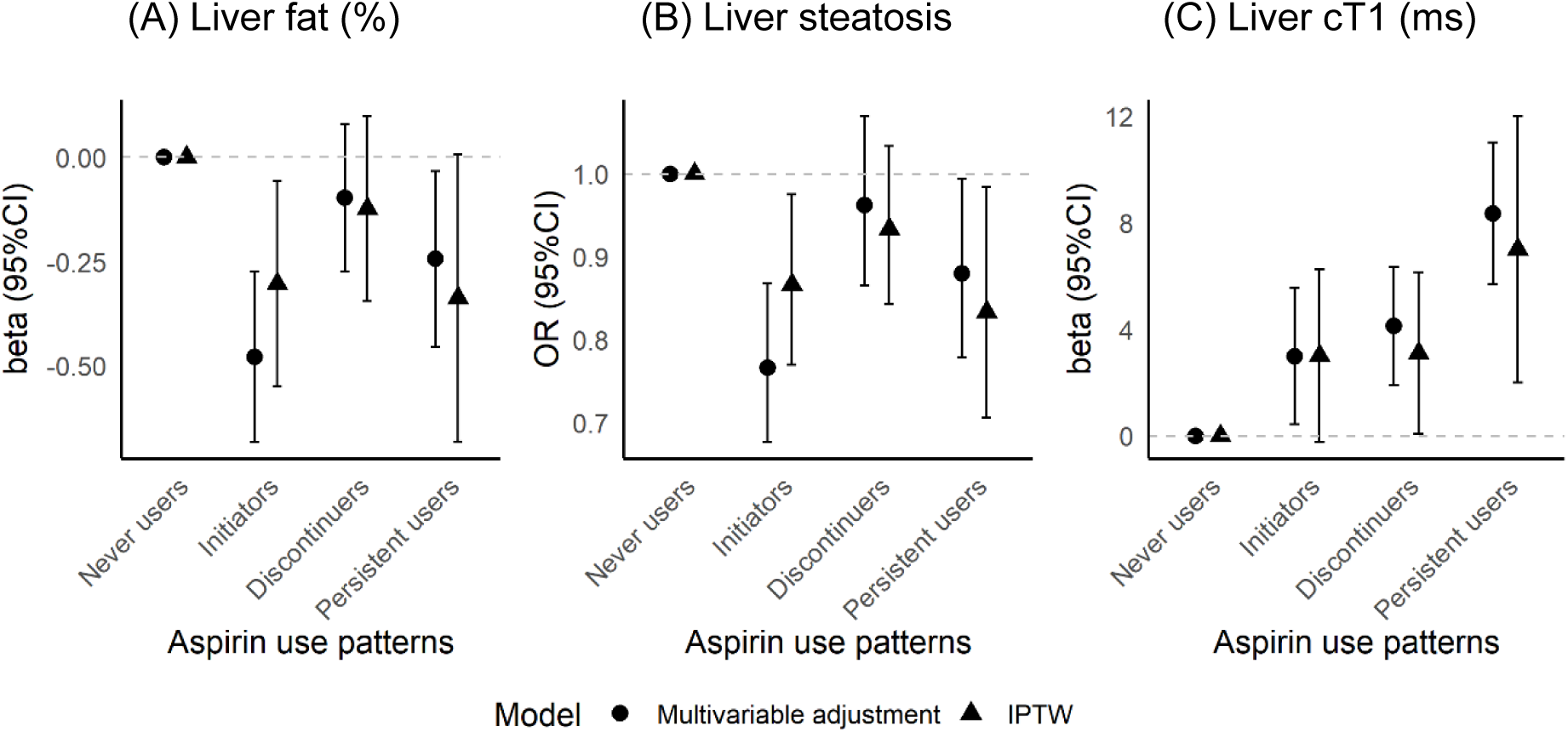
Associations between aspirin use patterns and liver fat, liver steatosis and cT1. Multivariable adjustment model included age, sex, deprivation index, ethnicity, physical activity, smoking, daily alcohol consumption, diabetes, hypertension, body mass index, high triglycerides, low high-density lipoprotein, existing cardiovascular disease, and baseline fatty liver index.

Subgroup analyses showed consistently negative associations between aspirin use at imaging visit and liver fat, and positive associations with liver cT1. Notably, the inverse association between aspirin use and liver fat was stronger among participants aged > 65 years compared to those < 65 years (-0.44 (-0.63, -0.26) vs. - 0.08 (-0.35, 0.19), p for interaction < 0.01). (supplementary table 2) Sensitivity analysis using truncated IPTW produced similar results to primary IPTW analysis (supplementary table 3).

## Discussion

In this cohort of 36413 participants, we found that both cross-sectional and longitudinal aspirin use were associated with lower liver fat and higher liver cT1. Multivariable adjustment and IPTW analyses showed consistent results, and the associations were robust across subgroups.

The observed inverse associations between aspirin use and liver fat percentage align with previous evidence. Lonardo and Zheng [13] reviewed epidemiological evidence supporting the antisteatotic effects of aspirin. Recent trial evidence has also shown low-dose aspirin reduced liver fat content, although in a small sample [16]. Shen et al [10] found that aspirin use was associated with lower NAFLD prevalence, especially in older adults, consistent with our findings of stronger associations in people > 65 years old. Using UKB data, Vell et al [27] showed similar findings that aspirin was associated with lower incidence of liver disease in men, although they did not examine quantitative liver fat or disease activity. However, Huang et al [14] found a positive association between aspirin and SLD incidence, though their study was limited by outcome definition of using only clinical diagnosis captured in medical records, introducing misclassification bias [17]. It is worth noting that the UKB imaging cohort had a mean liver fat of 4.9%, close to the diagnostic threshold for steatosis (≥5%), with only 27.8% of the participants exceeding this threshold. Although the observed aspirin-associated liver fat reduction was modest in absolute terms (0.35%), a shift of this magnitude could meaningfully alter the proportion of individuals classified as having steatosis at the population level, for example, to 30.1% in this cohort.

Several mechanisms may underlie the observed associations. Aspirin exerts both systematic anti-inflammatory effects and organ-specific effects, include promoting mitochondrial biogenesis via PGC-1α, attenuating hepatic collagen production through TGF-β1 inhibition, and suppressing platelet activation and pro-inflammatory signalling [13]. Preclinical studies show that platelets infiltrate the liver and promote inflammatory via Kupffer cells activation; whereas aspirin inhibits proinflammatory cyclooxygenase-2 and platelet derived growth factor signalling [16]. In animal models, aspirin reduced obesity and liver fat and improved glucose intolerance [28], and modulated the PPAR-AMPK-PGC-1α pathway in dyslipidemic states [29].

By contrast, we found that aspirin use was associated with increased liver cT1 by 5 ms, a biomarker of hepatic inflammation and fibrosis activity. This finding seems contradictory to some prior studies. Simon et al [11] and Jiang et al [12] reported that daily aspirin use was associated with reduced fibrosis in individuals with MASLD, assessed using non-invasive fibrosis scores, including fibrosis-4, NAFLD fibrosis score and AST/platelet ratio, although another study [15] found null associations. Importantly, however, the observed increase in cT1 in our study was minimal (5 ms), well below the clinically meaningful threshold of 80 ms [25]. Therefore, while statistically significant, this difference has limited clinical relevance and should not outweigh the potential antisteatotic benefits of aspirin.

Notably, many prior studies reporting protective effects of aspirin against fibrosis relied on blood-based surrogate scores (e.g., fibrosis 4, NAFLD fibrosis score, AST/platelet ratio), which largely reflect systemic biochemical correlates of liver injury and are sensitive to changes in hepatic necroinflammation and transaminase fluctuations. By contrast, cT1 captures local fibroinflammatory changes, odema and extracellular matrix, and may be less responsive to short-term metabolic improvements. Therefore, a plausible interpretation is that aspirin reduce hepatocellular fat accumulation and mitigate metabolic injury, but is less likely to rapidly reverse established imaging-detectable fibroinflammatory changes.

Alternative explanations remain possible. For example, aspirin users often have greater cardiometabolic burden, which may disproportionately influence cT1 and cause residual confounding. Temporal differences may also play a role. Antisteatotic effects of aspirin may occur earlier, while fibroinflammatory injury processes captured by cT1 progress over longer periods. Together, these findings suggest that aspirin reduces steatosis but does not necessarily rapidly protect against liver inflammation or fibrosis. Future investigations are needed to further examine the causal effects, with clear temporality, dose response relationship, and underlying mechanisms.

The strengths of this study include accurate quantitative measure of liver fat and cT1 via liver MRI, a large sample size, assessment of both cross-sectional and longitudinal aspirin use, and the application of multivariable adjustment and IPTW methods, which strengthen causal inference. However, there are also some limitations. First, aspirin use was self-reported as binary variable, without information on dose and duration, precluding dose-response analyses. Second, baseline liver MRI measures were unavailable, preventing direct evaluation of longitudinal changes in liver fat or cT1 over time; although we adjusted for baseline FLI, residual confounding from unmeasured liver changes may remain. Third, although we adjusted for a wide range of covariates, residual confounding cannot be entirely excluded. Application of multivariable adjustment and IPTW methods strengthened the result validity. Fourth, UKB data have restricted representativeness to general UK population, especially on ethnicity, health status and socioeconomic status, which may limit the generalizability of our findings to other populations. Finally, while MRI measures are highly informative, cT1 is an indirect measure of inflammation and fibrosis, and may not perfectly reflect liver fibroinflammatory injury.

## Conclusion

In summary, this large cohort study showed that aspirin use was associated with lower liver fat. The reduction in liver fat supports previous epidemiological and mechanistic evidence of aspirin’s potential antisteatotic effects, while the minimal increase in cT1 is not clinically meaningful. Future mechanistic and longitudinal studies with detailed exposure data, dose–response analyses, and direct measures of fibrosis progression are warranted to clarify aspirin’s role in the prevention and management of liver steatosis.

## Study highlights

### What is the current knowledge on the topic?

Aspirin has well-established cardioprotective and anti-inflammatory effects. Emerging evidence suggests it may influence liver fat, but its relationship with liver fat accumulation and fibroinflammation remains unclear. Non-invasive biomarkers such as MRI-derived proton density fat fraction (PDFF) and corrected T1 (cT1) provide precise assessment of liver steatosis and injury, yet few population-based studies have examined how aspirin use relates to these markers.

### What question did this study address?

This study investigated whether regular aspirin use and longitudinal use pattern are associated with liver fat (PDFF) and liver fibro-inflammation (cT1).

### What does this study add to our knowledge?

Among 36,413 UK Biobank participants, aspirin use was associated with lower liver fat but a small, clinically insignificant increase in cT1. Analyses of longitudinal aspirin patterns showed that initiators and persistent users had consistently lower liver fat than never users, while cT1 differences remained minimal. These findings suggest that aspirin may reduce liver fat through metabolic pathways without rapidly reversing fibroinflammatory injury.

### How might this change clinical pharmacology or translational science?

The results highlight aspirin’s potential role in modulating hepatic fat accumulation. While the modest cT1 increase indicates limited impact on fibroinflammation, these findings can inform mechanistic studies and guide the design of trials investigating aspirin as adjuncts for metabolic liver disease. Integrating real-world medication use with precise imaging biomarkers offers a robust framework for evaluating therapeutic repurposing in liver disease.

## Data Availability

All data produced in the present study are available upon reasonable request to the authors

## Statements

### Acknowledgement

UK Biobank has obtained Research Tissue Bank approval from its governing Research Ethics Committee, as recommended by the National Research Ethics Service. This research has been conducted using the UK Biobank Resource (application No. 74018). Permission to use the UK Biobank Resource was approved by the access subcommittee of the UK Biobank Board.

## Data sharing statement

UK Biobank data are available to registered researchers at https://www.ukbiobank.ac.uk/.

## Author contributions

QF conceived the research idea, performed primary analysis and drafted the manuscript. All authors conducted data analysis, interpreted results, critically reviewed and revised the manuscript.

